# On the assessment of more reliable COVID-19 infected number: the italian case

**DOI:** 10.1101/2020.03.25.20043562

**Authors:** Giuseppe Tradigo, Pietro Hiram Guzzi, Pierangelo Veltri

## Abstract

COVID-19 (SARS-CoV-2) is the most recent pandemic disease the world is currently managing. It started in China at the end of 2019, and it is diffusing throughout Italy, one of the most affected countries, and it is currently spreading through European countries and USA. Patients affected by COVID-19 are identified employing medical swabs applied mainly to (i) citizens with COVID-19 symptoms such as flu or high temperature, or (ii) citizens that had contacts with COVID-19 patients. A percentage of COVID-19 affected patients needs hospitalisation, whereas a portion needs to be treated in Intensive Care Units (ICUs).

Nevertheless, it is a matter of current intuition that COVID-19 infected citizens are more than those detected, and sometime the infection is detected too late. Thus there are many efforts in both tracking people activities as well as diffusing low cost reliable COVID-19 tests for early detection.

Starting from mortality rates of diseases caused by viruses in the same family (e.g. MERS, SARS, H1N1), we study the relations between the number of COVID-19 infections and the number of deaths, through Italian regions. We thus assess several infections being higher than the ones currently measured. We thus focus on the characterisation of the pandemic diffusion by estimating the infected number of patients versus the number of death. We use such an estimated number of infections, to foresee the effects of restriction actions adopted by governments to constrain virus diffusion. We finally think that our model can support the healthcare system to react when COVID-19 is increasing.

## 1 Introduction

The coronavirus disease COVID-19 was identified in Wuhan (China) on December 2019 [9]. The virus diffusion has been surprisingly rapid. Until now, COVID-19 has killed more people than SARS and MERS combined, despite the lower case fatality rate [8]. Due to the lack of vaccines and targeted therapies for preventing the diffusion [3], many governments adopted severe containment measures for minimising interactions among people and reducing their movements. To date the COVID-19 virus caused a total of 398, 107 confirmed cases around the world, of which 103, 334 recovers and 17, 453 deaths. In China 80, 981 have been the confirmed cases with more than 3, 000 deaths. Starting from mid-February, the virus diffused in the northern regions of Italy [12, 10]. The emergency is mainly due to severe illness by pneumonia requiring hospitalisation in Intensive Care Units (ICUs) with use of breathing ventilator supports. In [5] we studied the correlation among infections and ICU beds, to support strategies and measures for COVID-19.

At the time of this study, 23rd of March, COVID-19 is spreading in Italy and in more than 1 month an large number of both deaths and infections has been registered. Such numbers are diffused differently among the 20 Italian regions with different administration, economics and logistic background. Italian government adopted containment measures in mainly 4 different milestone dates as summarised in Table 2.

The percentage of death in China reports a ratio among death and infection of about 4.3%. Starting from the analysis of number of death with respect to number of identified infected in Italy, we map the percentage in Figure 1. The percentage follows an increasing law showing that the number of measured infected persons is probably underestimated.

**Figure 1:**
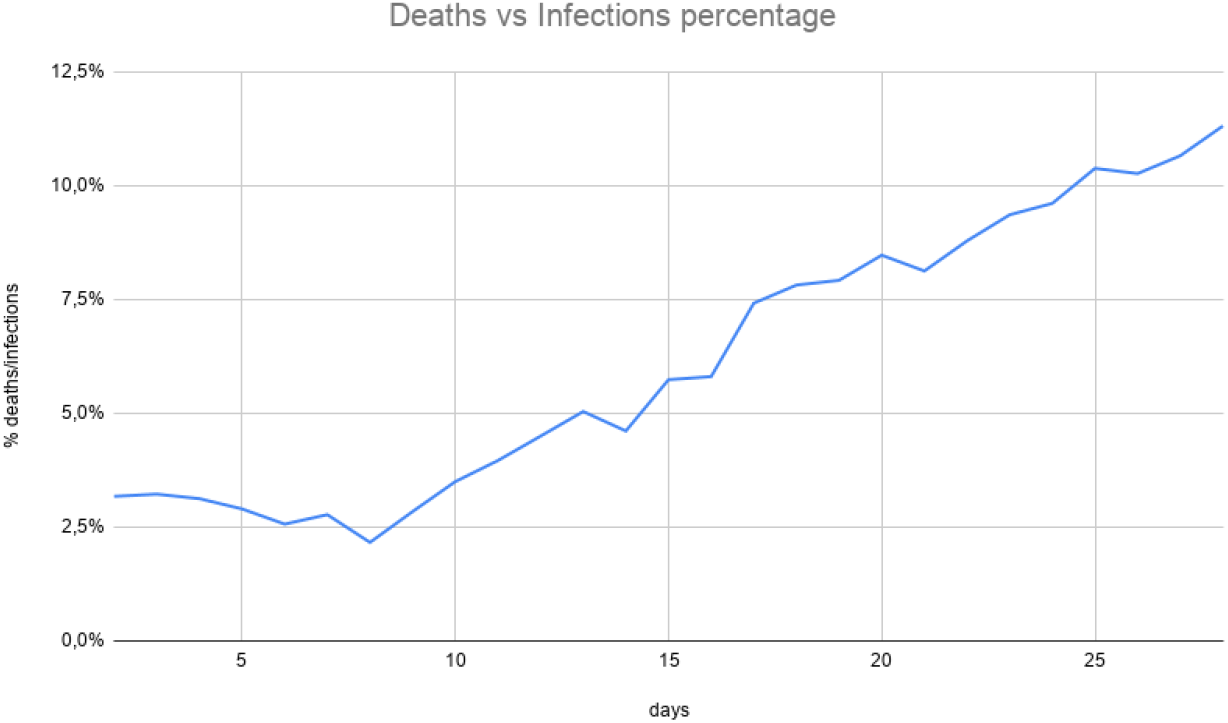
We report the percentage of deaths versus infections (i.e. CFR) in Italy. The value starts from acceptable rates but it reaches values above 11%, which is clearly not comparable with CFRs for similar viruses in the same family not with data from China or even world values estimated by the WHO.

The Coronaviridæ family contains many viruses, seven of which are known to be responsible for human diseases (229E, NL63, OC43, HKU1, MERS-CoV, SARS-CoV, and SARS-CoV-2) [7]. One of the main differences between the novel virus and the previous ones is its high spreading rate. Table1 reports the percentage of death known from Coronaviridæ family viruses exploited recently. We started observing that death percentage has an average value with respect to infected cases for all viruses. We report about hypotheses on a death percentage and we extract the number of infections. We compare such a number with respect to officially diffused infected ones. We finally use estimated numbers to evaluate containment measures with respect to infection diffusion.

## 2 Infections Number Estimation

We start from the analysis of epidemiological data from Wuhan city (China, Hubey region). As reported in [6, 4] about a third of infected patients became critical thus requiring ICU admission and breathing aids, due to severe pneumonia. The lesson learned from China has been used in other countries, such as Singapore and Italy, for preparing a correct strategy for emergency management.

In Italy ^1^ on March 22, we report a total of 59, 138 total cases (detected from 258, 402 swab tests), of which 46, 638 currently positives, 5, 476 deaths and 7, 024 recovered patients. Regarding infected people: 23, 783 are treated in their homes since they do not have severe ill, 22, 855 patients have been hospitalised, and 3, 009 patients have been admitted to ICUs.

At the date of March 23^*rd*^, while the situation in China seems to be now under control [11], the virus is continuing the diffusion in Italy and rapidly growing in other countries throughout the world [2]. Following Italy and China examples, other governments are implementing strict containment measures in order to dampen the spread of the infection. One of the main problems is related to the exponential diffusion of infections and also to the modality and protocols adopted for swab testing.

However, patients needing hospitalisation are fortunately a low percentage.

Nevertheless, in some cases, COVID-19 causes severe pneumonia, which requires respiratory support and can lead to death, especially in the presence of co-morbidities. Patients with severe pneumonia need to be treated in ICUs with the use of mechanical ventilators [1].

By analysing the Case Fatality Rates (CFR), we observe a value starting from low figures and increasing towards levels above 11%. Such a value seems not compatible with CFRs of other viruses of the same family and could be imputed to a bias in the choice of the swab test strategy. These tests are preferably performed on hub people, physicians, law enforcement agents and politicians and on people that have contact with infected ones. Some studies agree that COVID-19 CFR should be around 1 *−*2% and it is for sure below 4% [8].

Table 1 reports WHO’s CFR rate for COVID-19 being equal to 4.3%, which suffers the same problems we have just described. We claim that the total number of detected infections is much lower than the real ones due to the bias described above. We observe that we can derive the real cases of infections by exploiting the most reliable data available, which is the number of COVID-19 deaths. We can thus estimate the real infections by using the current number of deaths, starting by an hypothesis on the CFR value. We simulate three scenarios for CFR (1%, 2% and 3%) which are reported in Figure 2, in which we give an idea of the difference between measured data versus real once. The calculated values show 391.4% (*CFR* = 3%) more infections with respect to the measured ones, up to 1174% (*CFR* = 1%) in the worst scenario.

**Table 1:**
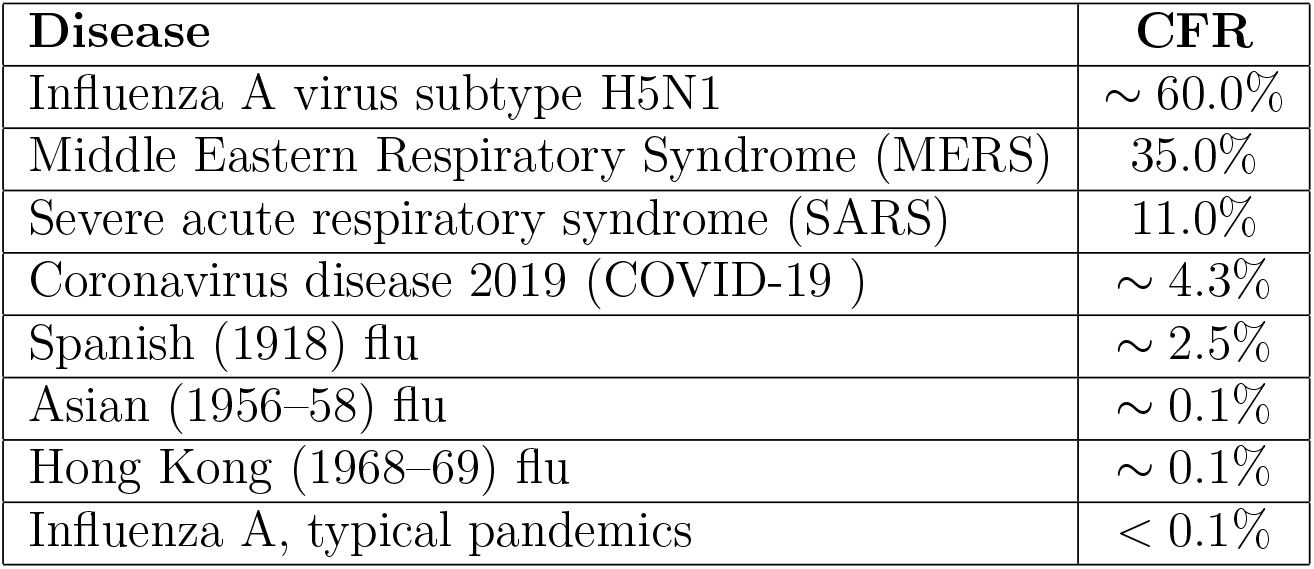
Case Fatality Rates (CFR) values for flu and diseases caused by Coronaviridiae virus. Even if COVID-19 virus shows a lower mortality rate, it killed more people than SARS-Cov1 (8098 cases and 774 deaths) and MERS-Cov (2494 cases and 858 deaths) combined. However we claim that the current rate of 4.3 is over estimated due to the bias in the swab strategy.

| Disease | CFR |
| --- | --- |
| Influenza A virus subtype H5N1 | $\sim 60.0\%$ |
| Middle Eastern Respiratory Syndrome (MERS) | 35.0% |
| Severe acute respiratory syndrome (SARS) | 11.0% |
| Coronavirus disease 2019 (COVID-19 ) | $\sim 4.3\%$ |
| Spanish (1918) flu | $\sim 2.5\%$ |
| Asian (1956–58) flu | $\sim 0.1\%$ |
| Hong Kong (1968–69) flu | $\sim 0.1\%$ |
| Influenza A, typical pandemics | $< 0.1\%$ |

**Table 2:** Containment measures and red zones definitions in Italy during COVID-19 emergency.

| Date | Event |
| --- | --- |
| March, 1<sup>st</sup> 2020 | first red zone in Codogno city (Lombardia region) |
| March, 8<sup>th</sup> 2020 | - red zone for the entire Lombardia region<br>- first containment measures for the whole country |
| March, 11<sup>th</sup> 2020 | red zone for whole Italy |
| March, 22<sup>nd</sup> 2020 | complete lockdown except for essential activities |

**Figure 2:**
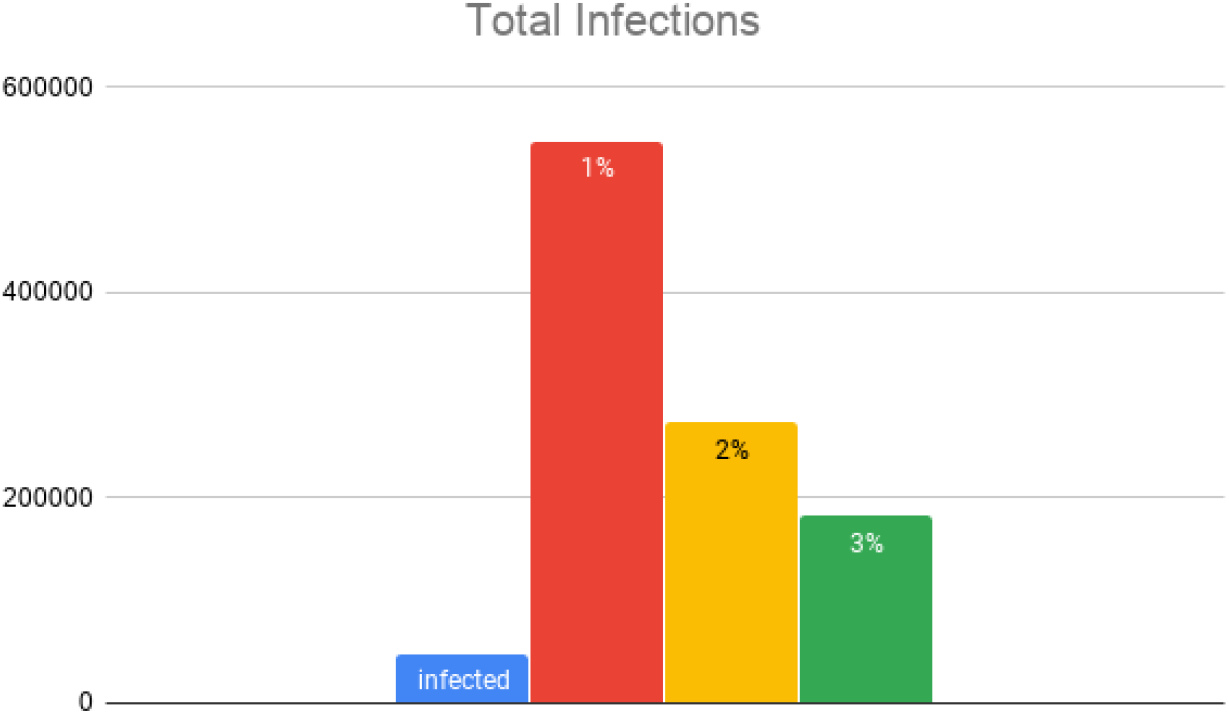
In blue the total infections currently detected in Italy via swab tests. The other bars report three scenarios with different Case Fatality Rates: *CFR* = 1% with 547, 600 infections (1174.1% more than currently detected), *CFR* = 2% with 273, 800 infections (587.1% more than currently detected) and *CFR* = 3% with 182, 533 infections (391.4% more than currently detected).

**Figure 3:**
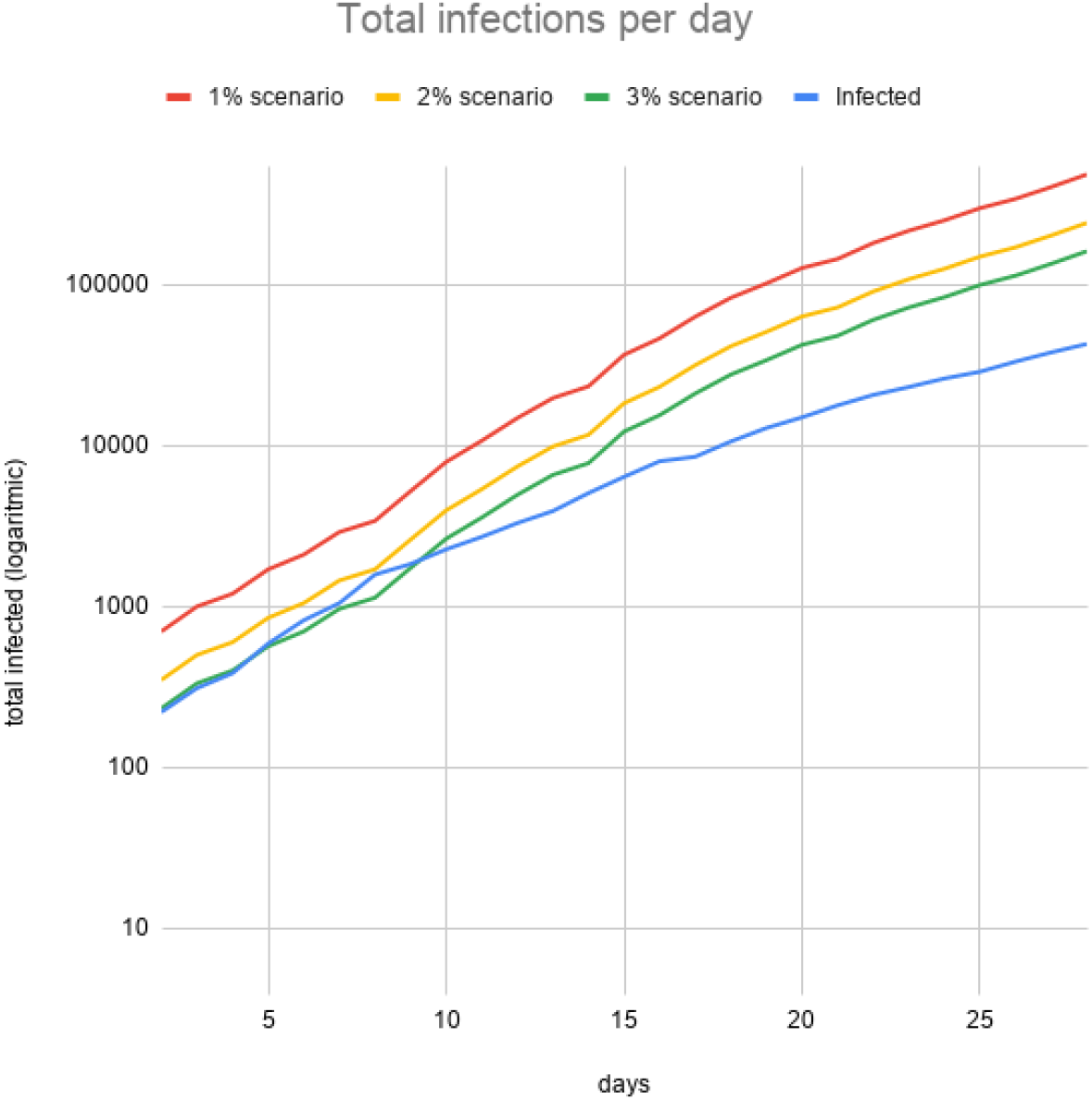
The figure reports infection levels in the three envisioned Case Fatality Rates scenarios (1%, 2% and 3%) compared to the current total COVID-19 infections detected via the swab tests. The curves are reported in the interval from February 24^*th*^ to March 22^*nd*^ 2020. Please note that the Y axis is logarithmic.

Such information can be useful for governments to plan actions for a better swab testing protocol and for ICU beds and resources strengthening at both a national and regional levels.

Finally, in Figure 2 we show the measured total infections (Y axis) per day (X axis) evaluated by using the above reported swab testing protocol and the scenarios of predicted infections for the three CFR values resulting in this study. The blue (i.e. lower) curve represents the number of recognised positive cases of the Italian official dataset.

## 3 Effects of Containment on COVID-19 diffusion

Governments are implementing containment plans to reduce the number of movements and also planning to track people movements to react to the exponential growth of infected patients. These measures has been implemented and universally applied to the whole population to reduce the probability of contacts in the hope of blocking the virus infections. The effects of such measures need several days for their effect to be seen. The most exposed people are the so-called hubs (or people having many social contacts), which have the highest probability of spreading the infection due to their jobs (e.g. law enforcement, physicians). For instance, the Chinese government applied severe mobility restrictions within the infected regions in order to block the virus infection [12].

We here report the effects of Italian containment measurements, by considering the main three events related to the red zones definitions, as reported in Table 2.

We plot the number of infections considering the first three above reported events. The last one (i.e. complete lockdown of March 22^*nd*^) is too recent to observe any impact on data. We plot such infections related to the number of performed swabs thus to have comparable infection trends in the three different time intervals: (i) day 0 to first containment event; (ii) from first containment to second one and (iii) from the second one to the extension of the red zone to the whole country.

As evidenced in Figure 4, even if effects could be identified in a time delay from any containment constraint, the upper trend has a slope that is lower than the other two ones. The second one can be considered with a marginal reaction with respect to the first one due to a delay in the application of containment measures.

**Figure 4:**
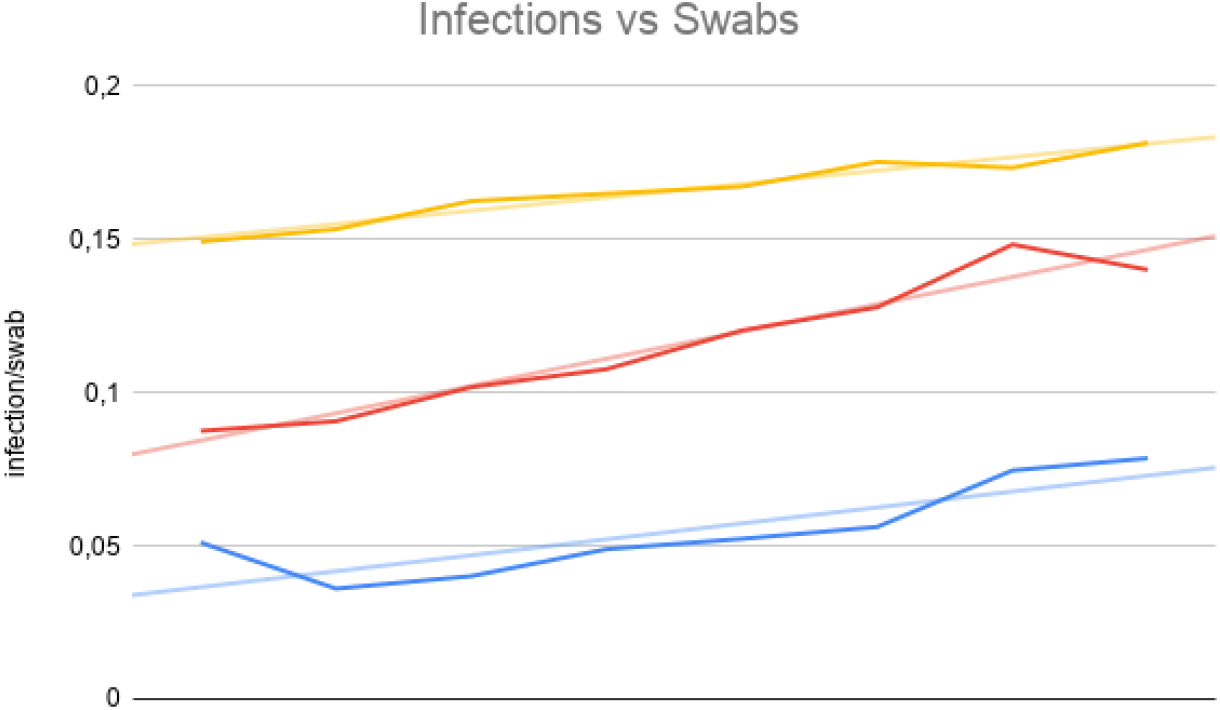
The figure reports infections versus swabs in three time windows: (*i*) before the first red zone of Codogno, (*ii*) during the definition of the red zone in the Lombardia region and (*iii*) after the red zone had been expanded to the whole country.

## 4 Scaling model at Regional distribution

Italy is divided in 20 regions that manage health structure in autonomy, under the central guidelines and funds. ICU beds resources as well as COVID-19 management and strategies follow central government guidelines. Nevertheless, regions and towns can be characterised by containment measurements. This allows a scalability in terms of rules and allows quarantine for large set of citizens. We here show that the above reported intuition and measurements are scalable also at regional and sub-regional scale.

We report the case of a large south region, where infections is growing up at with a time delay with respect to north Italian region (Lombardia) mostly due to workers and students moved from north to south of Italy after first containment measurements in north of Italy.

Figure 5 reports the infection rate of Lazio Region (central one) measured respect to number of death with different hypothesis of death rate as in Figure 2

**Figure 5:**
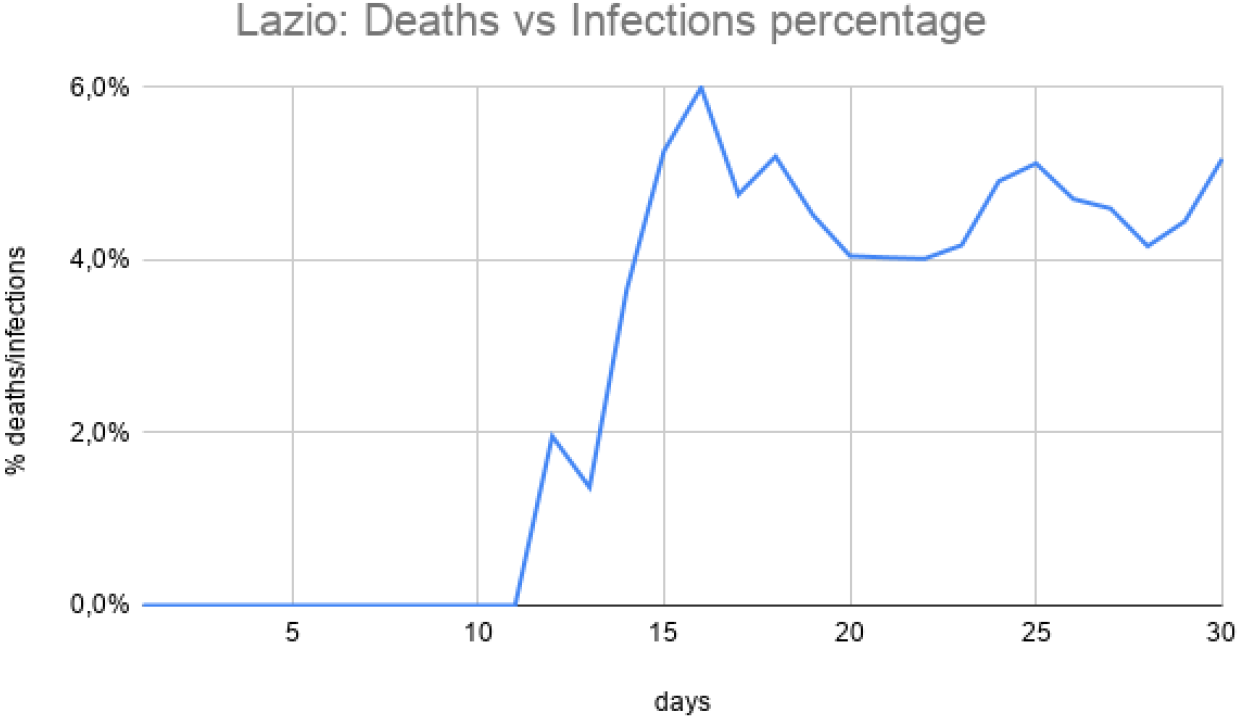
The figure depicts CFR values for the Lazio region. The value reaches figures equal to 6%, which is clearly related to the known bias about the currently adopted swab test protocol.

From such a Figure, the CFR value for the Lazio Region varies with respect to swab tests. If we consider the different CFR values reported above, the number of infected cases are plotted in Figure 6.

**Figure 6:**
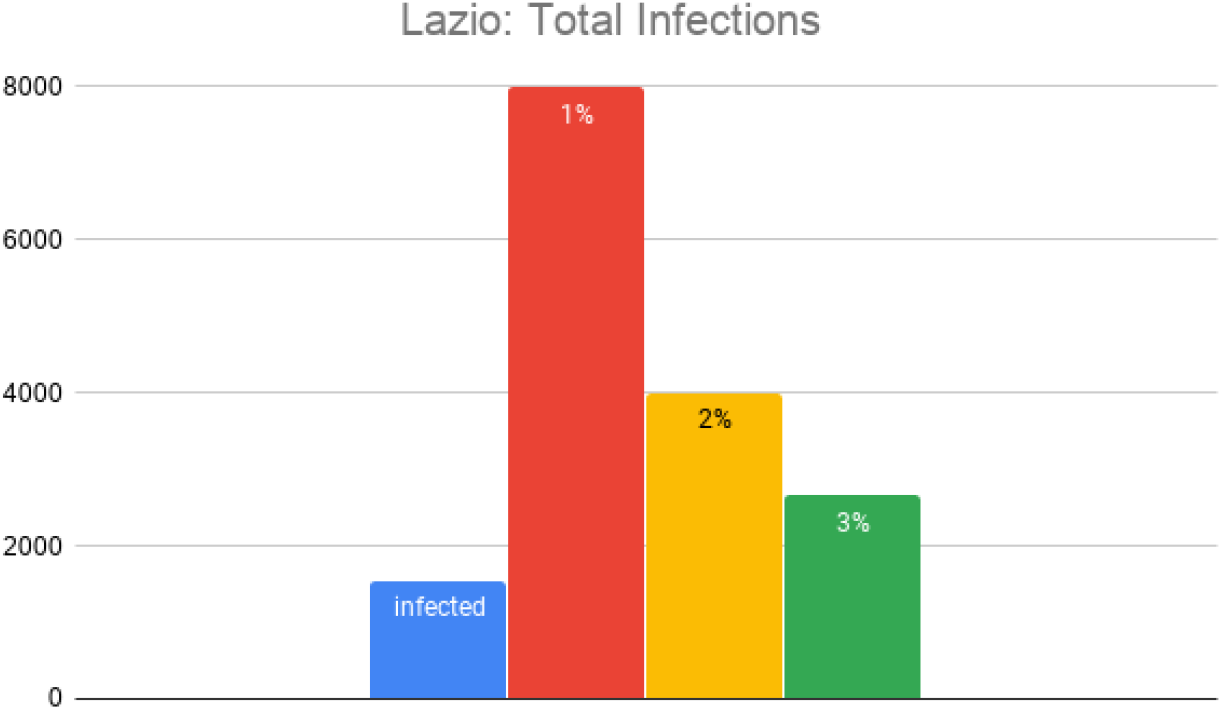
In blue the total infections (1, 545) currently detected in the Lazio region via swab tests. The other bars report three scenarios with different Case Fatality Rates: *CFR* = 1% (8, 000 infections, +517.8%), *CFR* = 2% (4, 000 infections, +258.9%) and *CFR* = 3% (2, 666 infections, +172.6%).

## 5 Conclusion

The emergency of COVID-19 is related to an aggressive virus that diffuses rapidly and strongly stresses the resistance of health structures. We started from observation of death percentage from previous coronavirus family viruses and we inferred the number of infections starting from CFR rates. The calculated infection levels should be considered to be more reliable than the current ones.

## Data Availability

All data are public available.

## 6 Contributors

GT was responsible for data analysis and statistics, and writing of the manuscript. PHG was responsible for data analysis and writing of the manuscript. PV was responsible for data analysis and writing of the manuscript. All the authors read and approved the manuscript.

## 8 Acknowledgments

We thank Italian Protezione Civile for freely providing online data thus allowing studies on COVID-19. We thank Prof. Gianluca Pollastri for his useful suggestions.

## Footnotes

1 http://www.salute.gov.it/portale/nuovocoronavirus/dettaglioContenutiNuovoCoronavirus.jsp

## References

[1] Yaseen M Arabi, Robert Fowler, and Frederick G Hayden. Critical care management of adults with community-acquired severe respiratory viral infection. Intensive Care Medicine, pages 1–14, 2020.

[2] Matteo Chinazzi, Jessica T. Davis, Marco Ajelli, Corrado Gioannini, Maria Litvinova, Stefano Merler, Ana Pastore y Piontti, Kunpeng Mu, Luca Rossi, Kaiyuan Sun, Cécile Viboud, Xinyue Xiong, Hongjie Yu, M. Elizabeth Halloran, Ira M. Longini, and Alessandro Vespignani. The effect of travel restrictions on the spread of the 2019 novel coronavirus (covid-19) outbreak. Science, 2020.

[3] Carlos del Rio and Preeti N Malani. Covid-19—new insights on a rapidly changing epidemic. Jama, 2020.

[4] Weijie Guan, Zheng-yi Ni, Yu Hu, Wenhua Liang, Chun-quan Ou, Jian-xing He, Lei Liu, Hong Shan, Chun-liang Lei, David SC Hui, et al. Clinical characteristics of coronavirus disease 2019 in china. New England Journal of Medicine, 2020.

[5] Pietro Hiram Guzzi, Giuseppe Tradigo, and Pierangelo Veltri. Intensive care unit resource planning during covid-19 emergency at the regional level: the italian case. medRxiv, 2020.

[6] Chaolin Huang, Yeming Wang, Xingwang Li, Lili Ren, Jianping Zhao, Yi Hu, Li Zhang, Guohui Fan, Jiuyang Xu, Xiaoying Gu, et al. Clinical features of patients infected with 2019 novel coronavirus in wuhan, china. The Lancet, 395(10223):497–506, 2020.

[7] Roujian Lu, Xiang Zhao, Juan Li, Peihua Niu, Bo Yang, Honglong Wu, Wenling Wang, Hao Song, Baoying Huang, Na Zhu, et al. Genomic characterisation and epidemiology of 2019 novel coronavirus: implications for virus origins and receptor binding. The Lancet, 395(10224):565–574, 2020.

[8] Elisabeth Mahase. Coronavirus: covid-19 has killed more people than sars and mers combined, despite lower case fatality rate, 2020.

[9] Monica Malta, Anne W Rimoin, and Steffanie A Strathdee. The coronavirus 2019-ncov epidemic: Is hindsight 20/20? EClinicalMedicine, 20, 2020.

[10] World Health Organization et al. Coronavirus disease 2019 (covid-19): situation report, 51. 2020.

[11] Fariba Rezaeetalab, Mahnaz Mozdourian, Mahnaz Amini, Zahra Javi-darabshahi, and Farzaneh Akbari. Covid-19: A new virus as a potential rapidly spreading in the worldwide. Journal of Cardio-Thoracic Medicine, 8(1):563–564, 2020.

[12] Joseph T Wu, Kathy Leung, and Gabriel M Leung. Nowcasting and forecasting the potential domestic and international spread of the 2019-ncov outbreak originating in wuhan, china: a modelling study. The Lancet, 395(10225):689–697, 2020.

